# Covid-19 Incidence And Mortality By Age Strata And Comorbidities In Mexico City: A Focus In The Pediatric Population

**DOI:** 10.1101/2021.06.21.21259282

**Authors:** Nadia González-García, María F Castilla-Peón, Fortino Solórzano Santos, Rodolfo Jiménez Juárez, Maria Elena Martínez Bustamante, Miguel Angel Minero Hibert, Juan Garduño-Espinosa

## Abstract

**Background:** SARS-COV2 appears less frequently and less severely in the pediatric population than in the older age groups. There is a need to precisely estimate the specific risks for each age group to design health and education policies suitable for each population

**Objective:** This study aimed to describe the risk of death in SARS-COV2 infected subjects by age group and according to the presence of comorbidities.

**Methods:** We analyzed data of confirmed SARS-COV2 infection cases where symptoms began between February 22th, 2020, and April 18th, 2021, as published by the General Epidemiology Direction (DGE) of the Mexican Ministry of Health. We calculated COVID-19 incidence and mortality by age group using population data from the Statistics and Population National Institute (INEGI), and estimated the association between risk of death and the presence of comorbidities.

**Results:** Mortality in SARS-COV2 infected people varied considerably, between 7 to 155 deaths per million per year in the under-20 age groups compared to 441 to 15,929 in the older age groups. Mortality in pediatric populations is strongly associated with comorbidities (OR: 4.6-47.9) compared to the milder association for older age groups (OR: 3.16-1.23).

**Conclusions:** The risk of death from SARS-COV2 infection in children is low and is strongly associated with comorbidities.

## Introduction

After more than a year, the COVID-19 pandemic has generated millions of hospitalizations and deaths worldwide. In addition to the direct impact caused by the SARS-COV2 infection itself, indirect harms have arisen because of the saturation of health systems, lockdown policies, and economic struggle.

Evidence indicates that children seldom develop a severe clinical presentation of SARS-COV2 infection and are less prone to transmit it than adults (1–4). For example, in China, more than 90% of infected children had a mild or moderate clinical presentation, a finding consistent with later reports (5,6). During the first three months of the pandemic, Mexico reported a 12.6% positivity rate for SARS-COV2 infection in tested children and a case-fatality rate of 1.9% (7). Mexico as a whole has been severely affected by the SARS-COV2 pandemic, as the overall death rate attests.

Despite the apparent low risk of severe disease in children, policies to mitigate the transmission rate in all age strata populations have restricted activities and settings essential for optimal child well-being and development. Therefore, accurate estimates of the health risks associated with SARS-COV2 infection in the pediatric population are necessary to design policies that optimize children’s well-being and development while protecting more susceptible groups.

This analysis aims to describe the epidemiology of SARS-COV2 in Mexico City, focusing on the children and adolescent population to appraise the specific health risks of this age stratum.

## Methods

We analyzed the epidemiological information for the SARS-COV2 pandemic in Mexico City. Data were extracted from the open-access data published by the General Epidemiology Direction (DGE) of the Mexican Ministry of Health, which reports data from all symptomatic and tested individuals (8). We included in the analysis the following three categories of cases: 1) virologically confirmed cases (positive RT-PCR or antigenic test) plus 2) confirmed cases by epidemiologic association (symptomatic contact of a virologically confirmed SARS-COV2 registered case in whom a valid nasal swab could not be performed), plus 3) cases without a valid virologic test confirmed by expert judgment. Deaths from any of these cases were recorded.

The DGE database includes information on the presence of specific comorbidities and risk factors, such as male sex, diabetes, immunosuppression, systemic hypertension, obesity, chronic renal disease, asthma, chronic obstructive pulmonary disease (COPD), tobacco use, or a report for “other comorbidity.” We calculated the frequency of each of these factors by age stratum and computed the risk of death in both the “comorbidity-free” population and the population having any of the reported comorbid condition by age-stratified groups.

The adjusted odds ratio (OR) for death in SARS-COV2 cases was calculated by logistic regression. We included factors significantly associated with death in the bivariate analysis.

We computed all-cause general mortality in 2019 and annualized COVID-19 mortality from February 22, 2020 to April 18th, 2021, using population data from The National Institute of Geography and Statistics (9)(10). We calculated the ratio of COVID-19 annualized mortality rate against 2019-all-cause mortality rate by age strata.

Analysis was performed using Stata software, version 13.0 (StataCorp), and graphs were made with GraphPad Prism version 9.1.0 for Windows (GraphPad Software).

## RESULTS

The DGE database includes 6,412,677 records of people tested for SARS-COV2 from February 20 to April 18,2021 (422 days), of which 629,527 were confirmed as COVID-19 cases by any of the following three means: 1) clinical-epidemiological association (n= 83,997, 13.3%), 2) by an expert committee judgment (n=2,281, 0.4%) or 3) a positive virological test (n=543,249, 86.3%). Overall positivity rates in those tested for SARS-COV2 were 34.5% and 19.7% for those tested with RT-PCR and antigen test, respectively.

Total cases, deaths, and frequency of comorbidities by age group are reported in Table 1. There were 52,432 total cases and 52 deaths in population younger than 20 years old. The significant differences in incidence and case-fatality rate between pediatric and adult age strata are noticeable, with a case-fatality rate below 0.3% in population between 1 and 20 years old which sharply increases beyond 40 years age and reaches more than 18% beyond 60. The frequency of specific registered comorbid conditions in the pediatric age group was very low, as most were coded as “other comorbidities.”

**Table 1.** Cases, deaths, and comorbidity frequencies by age group in Mexico City from February 22th to April 18th.

|  | <1<br>year | 1-4<br>years | 5-9<br>years | 10-14<br>years | 15-19<br>years | 20-39<br>years | 40-59<br>years | 60-79<br>years | 80-99<br>years |
| --- | --- | --- | --- | --- | --- | --- | --- | --- | --- |
| Total cases | 743 | 2,741 | 6,683 | 14,223 | 28,042 | 239,952 | 232,254 | 92,846 | 12,043 |
| Total deaths | 16 | 6 | 9 | 5 | 16 | 1,488 | 9,937 | 16,420 | 4,106 |
| Share of total deaths | 0.05% | 0.02% | 0.03% | 0.02% | 0.05% | 4.7% | 31.1% | 51.3% | 12.8% |
| Incidence per 1 million | 6,223 | 6,263 | 10,250 | 20,201 | 37,292 | 82,278 | 93,305 | 63,299 | 46,721 |
| Mortality per 1 million | 155 | 16 | 14 | 7 | 21 | 441 | 3,453 | 11,195 | 15,929 |
| Case-fatality rate (%) | 2.15 | 0.22 | 0.13 | 0.04 | 0.06 | 0.62 | 4.28 | 17.69 | 24.09 |
| COVID-19 / all cause 2019 mortality (%) | 1.4 | 2.8 | 5.9 | 3.0 | 2.5 | 29.6 | 62.1 | 90.8 | 86 |
| Male sex (%) | 53.6 | 52.1 | 51.7 | 50.2 | 49.6 | 48.3 | 47.4 | 49.9 | 47.3 |
| Indigenous n (%) <sup>1</sup> | 3 (0.4) | 9 (0.3) | 21 (0.3) | 48 (0.3) | 88 (0.3) | 1003 (0.4) | 1230 (0.5) | 585 (0.6) | 73 (0.6) |
| Diabetes n (%) | ND <sup>2</sup> | 4 (0.2) | 18 (0.3) | 52 (0.4) | 120 (0.4) | 4536 (1.9) | 30153 (13.0) | 26379 (28.5) | 3227 (26.9) |
| Inmunosupressed n(%) | 13 (1.8) | 19 (0.7) | 20 (0.3) | 42 (0.3) | 61 (0.2) | 805 (0.3) | 1698 (0.7) | 1262 (1.4) | 188 (1.6) |
| Systemic Hypertension n (%) | ND | 9 (0.3) | 8 (0.1) | 37 (0.3) | 117 (0.4) | 6152 (2.6) | 35543 (15.3) | 34649 (37.4) | 5847 (48.8) |
| Obesity n (%) | 48 (6.5) | 11 (0.4) | 117 (1.8) | 448 (3.2) | 1085 (3.8) | 22693 (9.5) | 31633 (13.7) | 11719 (12.7) | 1002 (8.4) |
| Chronic Renal Disease n(%) | 4 (0.5) | 3 (0.1) | 14 (0.2) | 19 (0.1) | 53 (0.2) | 899 (0.4) | 2038 (0.9) | 2200 (2.4) | 342 (2.9) |
| Chronic Obstructive Pulmonary Disease n(%) | 4 (0.5) | 2 (0.07) | 8 (0.1) | 10 (0.1) | 26 (0.1) | 289 (0.12) | 1272 (0.6) | 2365 (2.6) | 870 (7.3) |
| Asthma n(%) | 11 (1.5) | 33 (1.2) | 159 (2.4) | 403 (2.8) | 729 (2.6) | 5474 (2.3) | 4283 (1.8) | 1326 (1.4) | 144 (1.2) |
| Smoker n (%) | ND | ND | ND | 62 (0.44) | 1069 (3.82) | 29872 (12.5) | 22147 (9.6) | 7228 (7.8) | 807 (6.7) |
| Other comorbiditie(s) n (%) | 15 (2.0) | 36 (1.3) | 32 (0.5) | 72 (0.5) | 112 (0.4) | 1732 (0.7) | 2743 (1.2) | 2078 (2.3) | 467 (3.9) |
| 1 Includes all those who self-identified as indigenous or spoke an indigenous language |  |  |  |  |  |  |  |  |  |
| 2 No data |  |  |  |  |  |  |  |  |  |

Incidence and mortality by million people, case-fatality rate, and COVID-19/2019 all-cause-mortality ratio, are shown in Figure 1. The highest incidence of COVID-19 was observed in the 40-59 years old age group, with the case-fatality rate sharply increasing with age.COVID-19 mortality has surpassed more than 50% of the all-cause mortality in 2019 in age groups older than 40, while it has been less than 6% for those younger than 20.

**FIGURE 1.**
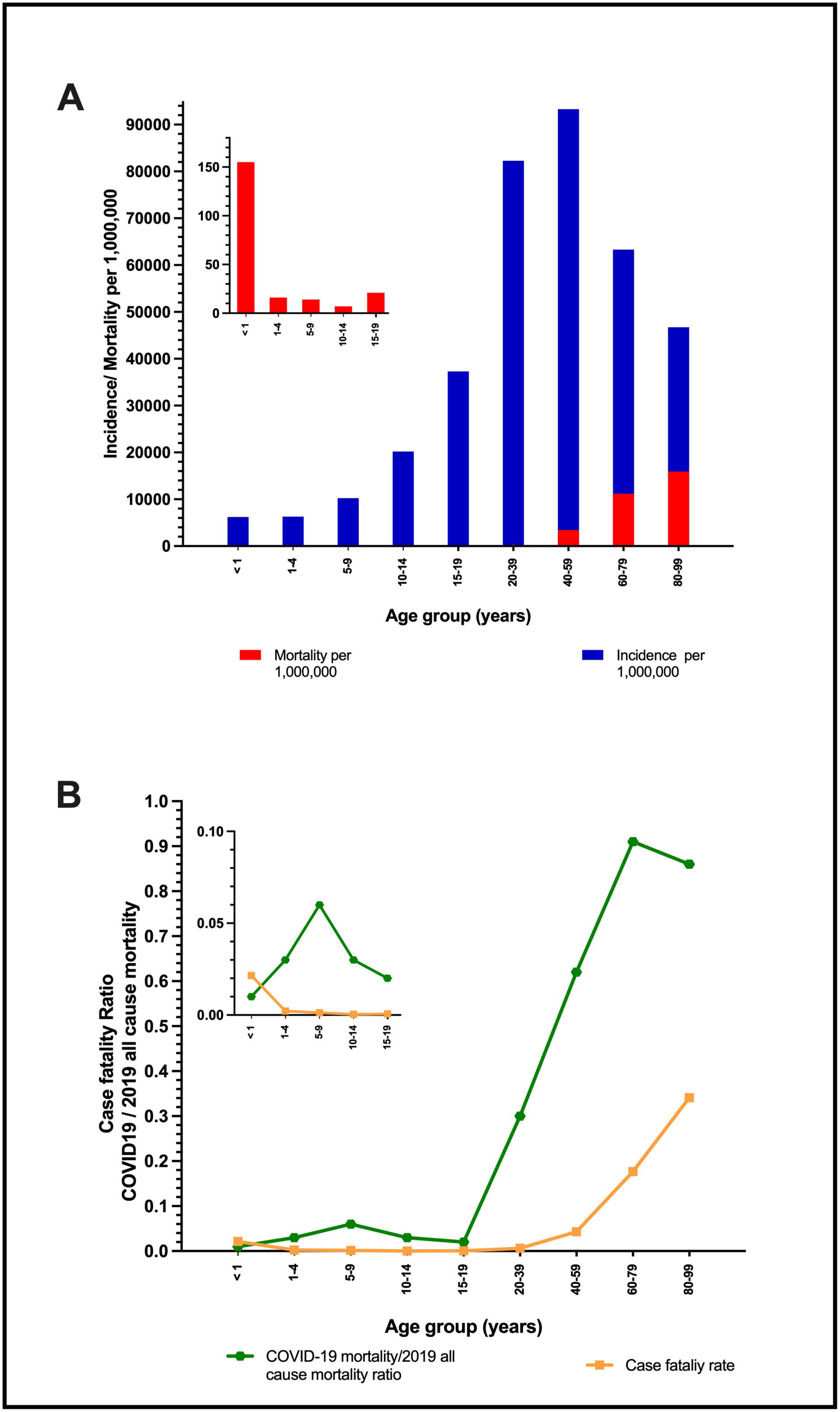
**A.** COVID-19 annualized incidence and mortality rates per 1,000,000 people **B.** case-fatality rate, and COVID-19/ 2019 all-cause mortality ratio by age strata

The adjusted OR for death in SARS-COV2 infected individuals for each comorbidity and risk factor reported in the database can be consulted in Table 2. Relative risk attributable to the presence of comorbidities was highest among children and adolescents, comorbidities being accountable for 80-98% of the age-specific mortality. In children under ten years of age, comorbid conditions associated with increased mortality were mainly those coded as “other comorbidities.” In those between 11 to 20 years old, diabetes, obesity, immunosuppression, and chronic renal disease were the most significant associated factors. Data for asthma, COPD, and tobacco use were not included because they had no statistical association with death risk in almost any stratum.

**Table 2.** Death risk in SARS-COV2 confirmed cases by comorbidities

| Table 2. Death risk by comorbidities in SARS-COV2 confirmed cases |  |  |  |  |  |  |  |  |  |  |  |
| --- | --- | --- | --- | --- | --- | --- | --- | --- | --- | --- | --- |
| N=629,<br>518 | Lethality if present any<br>comorbidity % (n) | Lethality if no reported<br>comorbidity %(n) | Relative Risk<br>for death if<br>comorbidity present<br>(IC95%) | Attributable risk relative<br>to comorbidity<sup>1</sup> | Adjusted Odds Ratio for Death |  |  |  |  |  |  |
|  |  |  |  |  | Male sex | Diabetes | Immunosuppression | Systemic<br>Hypertension | Obesity | Chronic Renal<br>Disease | Other comorbidities |
| <1<br>years | 5.8%<br>(9/154) | 1.2%<br>(7/589) | 4.9%<br>(1.9-13.0) | 79% | 5.3<sup>2</sup> | - | 3.2 | - | - | - | 5.4 |
| 1-4<br>years | 2.4 %<br>(3/127) | 0.1%<br>(3/2614) | 20.6%<br>(4.2-101.0) | 96% | 0.95 | - | - | - | - | - | 47.5<sup>3</sup> |
| 5-9<br>years | 0.5%<br>(2/378) | 0.1%<br>(7/6296) | 4.6%<br>(1.0-22.3) | 80% | 1.4 | - | - | - | 5.7 | 27.12<sup>2</sup> | 52.5<sup>3</sup> |
| 10-14<br>years | 0.4%<br>(4/1097) | 0.01%<br>(1/13126) | 47.9%<br>(5.4-427.8) | 98% | 2.9 | 111.2<sup>3</sup> | 6.6 | - | 1.93 | - | 141.7<sup>3</sup> |
| 15-19<br>years | 0.3%<br>(9/3121) | 0.03%<br>(7/24921) | 10.3%<br>(3.8-27.6) | 90% | 1.7 | 0.9 | 38.7<sup>3</sup> | 0.02 | 8.8<sup>3</sup> | 46.7<sup>3</sup> | 4.04 |
| 20-39<br>years | 1.3% (808/60904) | 0.4%<br>(680/179048) | 3.5%<br>(3.2-3.9) | 69% | 2.7<sup>3</sup> | 4.7<sup>3</sup> | 1.9<sup>2</sup> | 2.7<sup>3</sup> | 2.94<sup>3</sup> | 9.2<sup>3</sup> | 4.5<sup>3</sup> |
| 40-59<br>years | 6.6%<br>(6157/92828) | 2.7%<br>(3780/139426) | 2.5%<br>(2.4-2.5) | 59% | 2.7<sup>3</sup> | 2.4<sup>3</sup> | 1.8<sup>3</sup> | 1.7<sup>3</sup> | 1.93<sup>3</sup> | 4.89<sup>3</sup> | 3.0<sup>3</sup> |
| 60-79<br>years | 20.7%<br>(11234/54236) | 13.4%<br>(5186/38610) | 1.5%<br>(1.5-1.6) | 35% | 1.9<sup>3</sup> | 1.5<sup>3</sup> | 1.23<sup>2</sup> | 1.2<sup>3</sup> | 1.63<sup>3</sup> | 2.94<sup>3</sup> | 2.9<sup>3</sup> |
| 80-99<br>years | 36.5%<br>(2834/7763) | 29.7%<br>(1272/4280) | 1.2%<br>(1.2-1.3) | 19% | 1.8<sup>3</sup> | 1.2<sup>3</sup> | 1.2 | 1.2<sup>3</sup> | 1.44<sup>3</sup> | 1.46<sup>3</sup> | 2.2<sup>3</sup> |
| <sup>1</sup> (Lethality if comorbidity present – Lethality if no comorbidity) / Lethality if comorbidity present. <sup>2</sup>P <0.05, <sup>3</sup>P<0.001 |  |  |  |  |  |  |  |  |  |  |  |

## Discussion

This study describes the epidemiology of SARS-COV2 in the pediatric population in Mexico City as related to the epidemiology of other age strata. COVID-19 incidence and case-fatality rate in children younger than ten years are several times lower than in adults. Our results confirm those observed by others about the low frequency of the disease and the low rate of complications and deaths in this age group (5)(6). Besides, the case-fatality rate in youngsters is very strongly associated with present comorbidities.

The interpretation of these results is limited by the quality of the information provided by the DGA. Mexico is one of the countries with fewer SARS-COV2 tests relative to population (53.2 tests per 1000 people) (11), leading to a significant sub-estimation of cases. To increase the reliability of the information, we chose to limit the analysis to Mexico City since it is one of the Mexican entities with more comprehensive testing policies (283 tests per 1000) (12).

Besides, we decided to include cases confirmed by clinical-epidemiological association and expert panel judgment to mitigate the known underreporting derived from the low testing frequency. Together, non-virologically confirmed cases represent 14% of analyzed registers. While this decision might have biased absolute frequency estimates upwards, it does not modify our conclusion about the extremely low mortality by COVID-19 in the pediatric age group. Meanwhile, the relative estimates between age strata are still informative.

There is a lack of systematized information on the factors that increase the risk of death in the pediatric age group in Mexico. The mandatory epidemiological report in Mexico does not contain detailed information on the most frequent comorbidities in the pediatric age group, and those that exist frequently are reported under the umbrella term “other comorbidities.” This comorbidity category is the most commonly associated with lethality in the pediatric age group. Informal personal communications let us know that under this “umbrella term,” there is a high proportion of premature neonates with major congenital anomalies, disabled children with chronic neurological diseases, and cancer patients. A population-based cohort study in the United Kingdom(13) found 26,322 SARS-COV2 virologically confirmed cases younger than 18 years of age (1% cumulative incidence). Of these, 159 (0.006%) children were admitted to the hospital for more than 36 hours, while 73 were admitted to an intensive care unit (ICU). Comorbidities associated with hospital admission were type 1 diabetes (OR: 10.1, IC 95%: 4.12-24.8), congenital heart disease (OR: 2.69, IC 95%: 1.27-5.71), cerebral palsy (OR: 7.96, IC95%: 2.79 -22.76), epilepsy (OR: 6.17, IC 95%: 2.77-13.73) and sickle cell disease (OR: 8.24, IC 95%: 2.02-33.5). Interestingly, this study did not found a significant association between asthma and hospital admission, and neither did ours. Only one death during the study period was reported so that no analysis could be done for this outcome. Other comorbidities found to be associated with ICU admissions are obesity and prematurity (14)(15).

In addition to the diverse risk factors for death, there is a need to establish the causal role of SARS-Cov 2 in pediatric fatalities in our population, including whether the SARS-COV2 infection was an incidental finding in a severely ill child, a contributing agent in a vulnerable patient, a nosocomial infection in a person hospitalized for another severe disease or indeed a major cause of death. Besides, as was pointed in a previous report, the quality of health care might be an essential contributing factor for the increased case-fatality rate in comparison to other high-income countries (16).

While COVID-19 mortality represents a substantial proportion of the expected deaths in the pre-pandemic years or has almost equaled it in older age groups, there is no excess mortality rate in the pediatric age group. Relevant data on other health problems in the young Mexican population is helpful to contextualize the probability of death because of SARS-COV2 infection in children and adolescents (∼10 per million). For example, some of the preventable causes of death in Mexico City in 2019 were more frequent than COVID-19 mortality in pediatric age groups: congenital infections (1871 deaths per million in <1 age), violence (16 deaths per million in 1-4 year-olds), accidents (15 deaths per million in 5-14-year-olds) and suicide (47 deaths per million in 15-19-year-olds). Many other Mexican states had comparable or higher mortality because of malnutrition and gastrointestinal infections. (9,10) Besides deaths, additional important health issues are being affected as a secondary effect of the pandemic, such as immunization, (13) perinatal health care, and chronic diseases attention programs. (7)(14)(15) In Mexico, schools have been closed for about 50 million children and adolescents for longer than a year. Increasing evidence suggests that prolonged school closure, home confinement, and social restrictions could have significant consequences for the mental health of both children and adolescents. (8)(17)(18)

SARS-COV2 vaccination program started in December 2020. By the cutoff date for this analysis in April, less than 1% of the population had been fully vaccinated (11). Since then, this rate has been rapidly increasing, so the expected risks of SARS-COV2 infection in every age strata are expected to decline substantially.

## Conclusion

The risk of death from SARS-COV2 infection in children from Mexico City is low and is mainly associated with comorbidities. The low risk of direct health injury from SARS-COV2 infection in this age group should be balanced against other prevalent health risks exacerbated by the pandemic mitigating measures. More research should be done regarding the specific factors contributing to death in SARS-COV2 fatal cases to characterize better the vulnerable pediatric population and design policies targeted for them.

## Data Availability

All data are available at https://www.gob.mx/salud/documentos/datos-abiertos-152127

## References

1. Galow L, Haag L, Kahre E, Blankenburg J, Dalpke AH, Lück C, et al. Lower household transmission rates of SARS-CoV-2 from children compared to adults. J Infect [Internet]. 2021 Apr [cited 2021 May 18]; Available from: https://linkinghub.elsevier.com/retrieve/pii/S0163445321002097

2. Viner RM, Mytton OT, Bonell C, Melendez-Torres GJ, Ward J, Hudson L, et al. Susceptibility to SARS-CoV-2 Infection among Children and Adolescents Compared with Adults: A Systematic Review and Meta-Analysis [Internet]. Vol. 175, JAMA Pediatrics. American Medical Association; 2021 [cited 2021 May 18]. p. 143–56. Available from: https://pubmed.ncbi.nlm.nih.gov/32975552/

3. Zurl C, Eber E, Siegl A, Loeffler S, Stelzl E, Kessler HH, et al. Low Rate of SARS-CoV-2 Infections in Symptomatic Patients Attending a Pediatric Emergency Department. Front Pediatr. 2021 Apr;9:637167.

4. Galow L V., Haag L, Kahre E, Blankenburg J, Dalpke A, Lück C, et al. Lower Household Transmission Rates of SARS-CoV-2 from Children Compared to Adults - Results from the FamilyCoviDD19-Study. SSRN Electron J. 2021 Mar;

5. Children and COVID-19: State-Level Data Report [Internet]. [cited 2021 May 19]. Available from: https://services.aap.org/en/pages/2019-novel-coronavirus-covid-19-infections/children-and-covid-19-state-level-data-report/

6. Dong Y, Dong Y, Mo X, Hu Y, Qi X, Jiang F, et al. Epidemiology of COVID-19 among children in China [Internet]. Vol. 145, Pediatrics. American Academy of Pediatrics; 2020 [cited 2021 May 18]. Available from: https://pubmed.ncbi.nlm.nih.gov/32179660/

7. Rivas-Ruiz R, Roy-Garcia IA, Ureña-Wong K, Aguilar-Ituarte F, Vázquez-De Anda GF, Gutiérrez-Castrellón P, et al. Factors associated with death in children with COVID-19 in Mexico. Gac Med Mex. 2021;156(6):516–22.

8. Datos Abiertos Dirección General de Epidemiología | Secretaría de Salud | Gobierno | gob.mx [Internet]. [cited 2021 May 20]. Available from: https://www.gob.mx/salud/documentos/datos-abiertos-152127

9. Población total por entidad federativa y grupo quinquenal de edad según sexo, serie de años censales de 1990 a 2020 [Internet]. [cited 2021 May 20]. Available from: https://www.inegi.org.mx/app/tabulados/interactivos/?pxq=Poblacion_Poblacion_01_e60cd8cf-927f-4b94-823e-972457a12d4b

10. Principales causas [Internet]. [cited 2021 May 20]. Available from: https://www.inegi.org.mx/sistemas/olap/registros/vitales/mortalidad/tabulados/ConsultaMortalidad.asp

11. Ritchie H, Ortiz-Ospina E, Beltekian D, Mathieu E, Hasell J, Macdonald B, et al. Coronavirus Pandemic (COVID-19). Our World Data [Internet]. 2020 Mar 5 [cited 2021 Jun 18]; Available from: https://ourworldindata.org/coronavirus

12. Total de pruebas, total de positivos y tasa de positividad - Diccionario de Datos casos_positivos.xlsx - Portal de Datos Abiertos de la CDMX [Internet]. [cited 2021 Jun 18]. Available from: https://datos.cdmx.gob.mx/dataset/total-de-pruebas-total-de-positivos-y-tasa-de-positividad/resource/07a0cb5b-cfa3-48fa-bf66-fb879e7d4fcd

13. Saatci D, Ranger TA, Garriga C, Clift AK, Zaccardi F, Tan PS, et al. Association Between Race and COVID-19 Outcomes Among 2.6 Million Children in England. JAMA Pediatr. 2021;1–11.

14. Swann O V., Holden KA, Turtle L, Pollock L, Fairfield CJ, Drake TM, et al. Clinical characteristics of children and young people admitted to hospital with covid-19 in United Kingdom: Prospective multicentre observational cohort study. BMJ. 2020;370.

15. Kara A, Böncüoğlu E, Kıymet E, Arıkan K, Şahinkaya Ş, Düzgöl M, et al. Evaluation of predictors of severe-moderate COVID-19 infections at children: a review of 292 children. J Med Virol. 2021 Jul;

16. Agulló V, Fernández-González M, Ortiz de la Tabla V, Gonzalo-Jiménez N, García JA, Masiá M, et al. Evaluation of the rapid antigen test Panbio COVID-19 in saliva and nasal swabs in a population-based point-of-care study. Vol. 82, Journal of Infection. 2021. p. 186–230.

17. Roberton T, Carter ED, Chou VB, Stegmuller AR, Jackson BD, Tam Y, et al. Early estimates of the indirect effects of the COVID-19 pandemic on maternal and child mortality in low-income and middle-income countries: a modelling study. Lancet Glob Heal. 2020;8(7):e901–8.

18. Sevilla A, Phimister A, Krutikova S, Kraftman L, Farquharson C, Costa Dias M, et al. Learning during the lockdown: real-time data on children’s experiences during home learning. 2020 May.

